# Micronutrient deficiency among pregnant adolescents in South Asia: A Systematic Review

**DOI:** 10.1101/2023.10.12.23296939

**Authors:** Blessing Akombi-Inyang, Mansi Dhami, Judith Byaruhanga, Zohra S. Lassi, Kingsley Agho

## Abstract

**Objectives:** Micronutrient deficiency is an important global health concern with great impact on growth and development outcomes, which may lead to substantial losses in overall productivity and potential. The burden of micronutrient deficiency negatively impacts the health of vulnerable groups, including pregnant adolescents. Hence, this study will systematically review the available evidence on micronutrient deficiency among pregnant adolescents in South Asia.

**Methods:** This systematic review adhered to the 2015 Preferred Re-porting Items for Systematic reviews and Meta-Analysis (PRISMA) guidelines. A combination of selected keywords was used to search 8 computerized biblio-graphic databases: Scopus, Ovid MEDLINE, PUBMED, EMBASE, PsycINFO, CINAHL, ProQuest, and Web of Science. Potential studies were imported into an Endnote library and screened for eligibility using pre-determined criteria. The quality of the included studies was assessed using the National Institutes of Health (NIH) checklist.

**Result:** There is a dearth of studies on micronutrient deficiency among pregnant adolescents in South Asia. Of a total of 616 studies, five studies met the inclusion criteria. The factors associated with micronutrient deficiency among pregnant adolescents in South Asia were food (in)security, intake of dairy products, low maternal knowledge, and inadequate food intake.

**Conclusion:** Our review suggested that lack of awareness and knowledge of adequate micronutrient intake and household food insecurity were associated with micronutrient deficiency among pregnant adolescents. Hence, interventions targeting pregnant adolescents are needed in South Asia and such interventions could include cash transfers integrated with nutrition and health interventions.

## Introduction

Micronutrients are an important component of maternal nutrition and are essential for biologic activities. They comprise of essential minerals and vitamins mainly obtained from the diet for the sustenance of key cellular and molecular functions [1]. The role of micronutrients in disease prevention, human development, and well-being, including the regulation of metabolism, bone density, heartbeat, and cellular pH is quite important. Though needed in minute quantity, consuming the recommended amount is important at every stage of life. Micronutrient status varies widely throughout pregnancy and across populations. Of all micronutrients, six have been identified as essential including iron, vitamin A, vitamin D, iodine, folate, and zinc [2]. Critical micronutrients in pregnancy include folate, calcium, iron, zinc, and iodine. Folate insufficiency increases the risk of neural tube defects (NTD) while excessive intake of vitamin A may affect the normal development of an embryo or foetus [3]. Deficiencies in iodine, vitamin A and iron are the most common globally, particularly in pregnant women [3]. In low- and middle-income countries (LMICs), women often commence their pregnancy journey malnourished, and the demand of gestation exacerbates existing micronutrient deficiencies and increases their susceptibility to infections, with health consequences for the foetus [3, 4].

Due to a phase of rapid somatic growth for their bodies and for the foetus they are bearing, pregnant adolescent girls (much more than their adult counterparts) are susceptible to micronutrient deficiency [5 – 6]. The health and social complexities of the adolescent’s transition to adulthood are far-reaching and well-documented [7, 8], but this is of-ten overlooked by family members and healthcare providers alike, accounting for why nutritional deficiencies is often missed [9]. The social change adolescents encounter often leads to unwanted pregnancies, which worsens an existing malnourished state [10, 11].

Approximately 12 million girls aged 15–19 years and at least 777,000 girls under 15 years give birth each year in developing regions [12, 13]. Pregnancy during adolescence poses significant risks of morbidity and mortality for both the mother and the baby, which may be exacerbated by micronutrient deficiency [14, 15]. Maternal nutritional status before and during early pregnancy is an important determinant of child health, especially among pregnant adolescents.

South Asia is home to one of the largest number of people with malnutrition globally [16]. Countries in South Asia face widespread hunger and micronutrient deficiency which leads to challenges in population health. Pregnant women and children are most susceptible to micronutrient deficiencies due to higher physiological requirements during pregnancy and childhood development which often lead to in-creased demand for specific vitamins and minerals.

Despite the importance of micronutrients to pregnant adolescents, there is a dearth of evidence on the underlying factors that affect the micronutrient status of pregnant adolescents, especially in South Asia. Considering the similar sociocultural characteristics of populations across the region, there is a need to account for contextual factors that may exist which require a global health approach. Given the association between micronutrient deficiency and the wellbeing of pregnant adolescents as well as the growth and development of their foetus [17] we are curious about available evidence which could possibly inform the design of a tailored nutritional intervention to improve the health outcome of this vulnerable subgroup. This study reviews factors associated with micronutrient deficiency in pregnant adolescents in South Asia, drawing on evidence from previous studies and discussing the need for more robustly designed studies on micronutrient deficiency in pregnant adolescents and provide a stronger evidence base against which future nutritional interventions can be developed.

## Methods

This systematic review was conducted in accordance with the Preferred Reporting Items for Systematic Reviews and Meta-Analyses (PRISMA) guidelines (S1). The PRISMA statement is a detailed checklist with 27 essential items for ensuring reporting transparency [18]. To avoid duplication, Google scholar and Cochrane library were initially searched to ensure no systematic reviews or meta-analyses on micro-nutrient deficiency among pregnant adolescents in South Asia had been previously conducted.

This systematic review is registered in PROSPERO international prospective registry for systematic reviews with reference CRD42020173325.

### Outcome variable

The outcome variable for this review is adolescent pregnancy, defined as pregnancy in women aged 10-19 years [19].

### Exploratory variable

Micronutrient deficiencies, which is defined as a lack of essential vitamins and minerals required in small amounts by the body for proper growth and development. They include (but not limited to) iron, vitamin A, calcium, vitamin D, iodine, folate, and zinc [1, 2].

### Search strategy

Following discussion of the research question among the authors, a list of relevant keywords was generated and used to comprehensively search for peer-reviewed articles from eight computerized biblio-graphic databases: Scopus, Ovid Medline, Embase, CINAHL, PubMed, ProQuest, PsycINFO, Web of Science. The following combination of keywords was used in the search:

(Micronutrien* OR vitamin A OR vitamin D OR iron OR zinc OR iodine)

AND

(Pregnant adolescent* OR pregnant teenager OR adolescent pregnanc* OR teenage pregnanc*)

AND

(India OR Pakistan OR Bangladesh OR Sri Lanka OR Nepal OR Afghanistan OR Bhutan OR Maldives OR South Asia)

The search was restricted to only studies conducted in South Asia with no publication year limitation. The studies retrieved from each database were exported into an EndNote library. In the case of relevant publications which might have been missed during the search, the bibliographical references of all retrieved studies that met the eligibility criteria were also searched and complemented by citation tracking using Google Scholar.

### Inclusion and Exclusion Criteria

We included studies that focused on: (i) pregnant adolescents aged 10 – 19 years; (ii) were conducted in South Asia (Afghanistan, India, Pakistan, Bangladesh, Sri Lanka, Nepal, Bhutan and Maldives); (iii) analyzed factors associated with micronutrient deficiency; (iv) were published in a peer-reviewed journal and their full texts were available; (v) were written in English and accessible as we did not have the logistical and financial capacity to translate literature published in languages other than English.

Studies were excluded if they focused on: (i) non-pregnant adolescents or pregnant/non-pregnant adult women; (ii) conducted in countries outside South Asia; (iii) published in languages other than English; (iv) were non-peer-reviewed research, reviews, commentaries, editorials, letters to editors, and opinion pieces; (v) and/or did not assess the factors associated micronutrient deficiency.

### Data extraction

The retrieved studies from the search of the databases were exported into an EndNote library, and all duplicates were removed. The exported articles were first screened by title and abstract. Thereafter, their full texts were assessed against the eligibility criteria. All aspects of data screening, extraction, and appraisals of identified studies were carried out by two independent reviewers (BA and MD), and all disagreements between the two reviewers were resolved through discussion and consensus. Where uncertainty emerged, a third reviewer (KA) was consulted to adjudicate differences in the selection of the final studies for inclusion.

A piloted form adapted from the Cochrane Handbook for Systematic Reviews [20] was used in the extraction of data from eligible studies. Data extracted include author, year of publication, country, study design, sample characteristics, micronutrient/deficient micronutrient, factors associated with micronutrient deficiency and quality assessment score (Table 1).

**Table 1:** Characteristics of included studies.

| Author [Ref]<br>Year, Country |  | Study design | Study aim | Sample Characteristics<br>(N = Number of participants) | Micronutrient/<br>Micronutrient Deficiency | Factors associated with Micronutrient Deficiency | Quality Assessment<br>[0 -10 points] |
| --- | --- | --- | --- | --- | --- | --- | --- |
| Rang-Din Nutrition Study [21, 22]<br>Bangladesh | Dewey et al. [21]<br>2019 | Cross-sectional study | To examine the effects of prenatal nutritional supplementation among pregnant adolescents and to explore whether the effects of the intervention differed by household food security | N = 1552<br>Average age: $17.4 \pm 1.3$ years<br>Average gestational age: 13 weeks | Not specified | Food insecurity | 7 |
| | Mridha et al. [22]<br>2018 | Cross-sectional study | To describe the nutritional and dietary characteristics of rural Bangladeshi adolescents during early pregnancy and identify factors associated with their nutritional status and dietary practices | N = 1552<br>Average age = $17.4 \pm 1.3$ years<br>Average gestational age = $13.0 \pm 3.4$ weeks | Iron Deficiency Anaemia | Higher intake of dairy products | 7 |
| Bano F. [23]<br>2019<br>India |  | Cross-sectional study | To estimate the prevalence of anaemia among pregnant women and its association with the various socio-demographic determinants in urban areas of Kanpur. | N = 40<br>Age = <20 years | Iron Deficiency Anaemia | Inadequate iron-folic acid supplementation, lack of antenatal registration | 6 |
| Nguyen et al. [24]<br>2017<br>BangladeshV |  | Cross-sectional study | To compare maternal and child nutrition and health conditions of pregnant adolescents with those of non-adolescents | N = 404<br>Age = 13–19 years | Calcium Deficiency | Low maternal knowledge | 7 |
| Pathak et al. [25]<br>2003<br>India | | Cross-sectional study | To assess the prevalence of iron, vitamin A and iodine deficiencies amongst rural Adolescent Pregnant Mothers | N = 151<br>Average age = $17.8 \pm 1.5$ years<br>Gestational age = $\geq 24$ weeks | Iron Deficiency Anaemia and Vitamin A<sub>1</sub> Deficiency | Inadequate dietary intake of micronutrients | 7 |

### Quality Assessment

Quality assessment of studies involves considering the risk of potential for selection bias, information bias, measurement bias, or confounding. The quality of the retrieved studies was assessed using the National Institutes of Health (NIH) checklist [26]. This checklist considers 14 distinct criteria in assessing the internal validity of studies. Scores apportioned to each included study range from 0 [if none of the criteria were met] to 14 points [if all criteria were met]. The overall quality of a study is dependent on the sum of points awarded. Studies rated as poor quality had a score <4, medium quality 5 – 9, while high quality studies had a score between 10 and 14, as minimal risk of bias and vice versa [26]. In addition, the more emphasis in the study design to issues around the establishment of a causal relationship between the expo-sure and outcome, the higher the quality of the study. Emerging evidence suggests that the NIH checklist is a robust tool for assessing risk bias in controlled intervention studies and cross-sectional studies [26-29].

## Results

The initial search yielded 616 studies from eight databases. After the removal of duplicates, 554 studies were retained. Screening by title and abstract resulted in the exclusion of 543 studies. The full texts of the remaining 11 studies were retrieved and read for eligibility and relevance, and nine studies were further excluded. Two studies met the eligibility criteria. A further manual search of the reference lists of the eligible studies identified two additional studies, resulting in a total of 5 studies included in this review, as shown in Figure 1.

**Figure 1.**
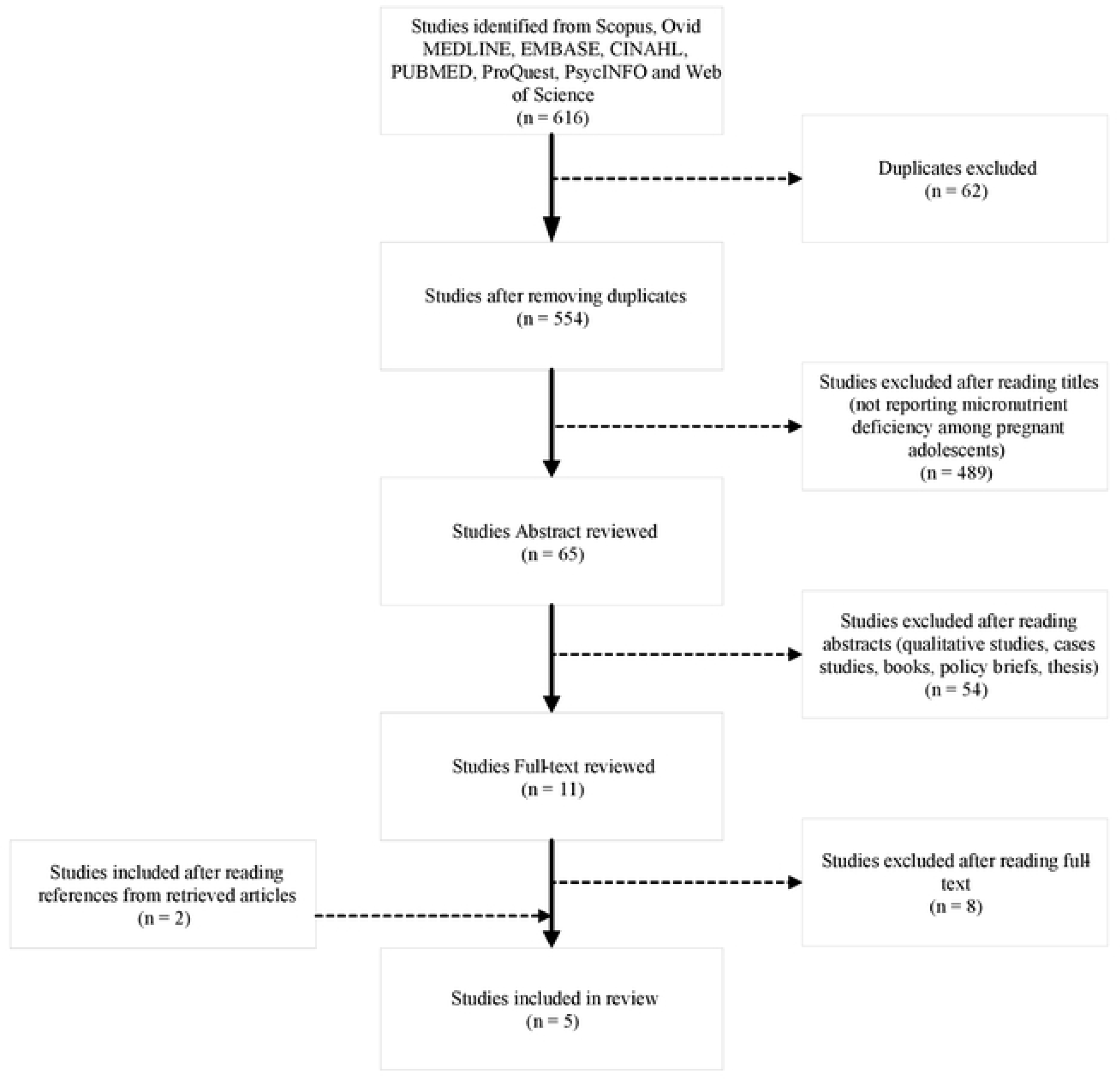
PRISMA flowchart for the selection of eligible studies.

### Characteristics of Included Studies

Table 1 shows a summary of the studies included in this review. Three studies were conducted in Bangladesh, and two in India. The sample size in the retained studies varied from 151 to 2000 participants, while the age was between 13 to 19 years. The 14 criteria used to evaluate the quality of included studies showed that all four studies were of medium quality. The details of the domain-specific score are provided in Supplementary Table S2.

## Discussion

A significant proportion of adolescent girls suffer from micronutrient deficiency, which may be made worse by pregnancy. Our finding suggests a dearth of studies on micronutrient deficiency among pregnant adolescents in South Asia. Of the studies re-viewed, the factors associated with micronutrient deficiency among pregnant adolescents in South Asia were food (in)security, advancing age, inadequate iron-folic acid supplementation, lack of antenatal registration, intake of dairy products, low maternal knowledge, and inadequate intake of food.

### Food (in)security

Dewey et al. [21] reported that women in food-insecure households are more likely to suffer from both macro- and micronutrient deficiencies during pregnancy, and this was evident in the subset of pregnant adolescents in the study. Inadequate intake of micronutrients in food insecure households could result from under-consumption of food or over-consumption of an energy-dense, but nutrient-poor diet. This might be more pronounced among pregnant adolescents due to lack of financial autonomy and knowledge to make more nutrition informed decision. Dewey et al. [21] also observed that food insecurity prevents pregnant adolescents from responding to nutrition supplementation interventions during the prenatal period. The uptake of these interventions is particularly important for pregnant adolescents with suboptimal micronutrient intake, as these interventions could serve as effective solutions to micronutrient deficiency in pregnancy. Furthermore, evidence has shown that food insecurity adversely affects women throughout their reproductive life from preconception [30] through to the postnatal period, with the pregnant adolescent being the most impacted [31]. A study conducted in Bangladesh reported that household food insecurity is a significant predictor of iron deficiency anaemia among women of reproductive age [32]. The finding from the Bangladeshi study was buttressed by other studies conduct-ed in Brazil and the United States, which also reported a chance of 2.63 and 3 times higher of pregnant women developing iron deficiency anaemia in food insecurity compared with those in situations of food security, respectively. The findings from these studies could be extrapolated to food-insecure pregnant adolescent women in South Asia due to their greater propensity towards micronutrient deficiency. In addition, a recent multicounty survey showed that the likelihood of anaemia is significantly higher among younger women [33]. Among other reasons, iron and folic acid deficiencies were the leading cause.

### Intake of dairy products

Dairy products are not only deficient in Vitamin C which is one of the main enhancers of iron absorption, but they are also very low in iron content and inhibit the absorption of iron present in other foods, for instance, calcium and phosphates in milk form insoluble complexes with iron. In their study, Mridha et al. [22] reported that iron deficiency is associated with a higher intake of dairy products. Milk and other dairy products can interfere with the gut’s ability to absorb iron from other sources, such as meat, meat alternatives, and dark green vegetables. One possible mechanism is the inhibition of non-heme iron absorption by calcium and casein present in dairy products [34]. However, Mridha et al. [22] ruled out this mechanism because only 10% of the study population exhibited iron deficiency. Though research has shown that infant and toddler consumption of dairy products, especially cow’s milk, has adverse effects on their iron stores, no conclusive evidence has been found on the association between the intake of dairy products and micronutrient deficiency in pregnant adolescents.

### Lack of antenatal care

Bano F. [23] reported a lack of antenatal registration among pregnant adolescent girls. Young girls are at increased risk of maternal and foetal complications due to in-adequate antenatal care and appropriate nutrition. Antenatal care is essential for the health and wellbeing of both the mother and the newborn. Adequate antenatal care can help identify and manage potential pregnancy complications, including pre-eclampsia, gestational diabetes, and other medical conditions. It also provides an opportunity for healthcare providers to educate mothers on healthy practices such as good nutrition, exercise, and the importance of prenatal vitamins. The lack of antenatal visits in adolescent pregnant women can increase the risk of pregnancy-related complications and adverse birth outcomes. An antenatal visit provides an opportunity to screen pregnant women for micronutrient deficiencies and provide them with appropriate supplements, monitor the foetus’s growth and development, intervene with appropriate supplements, and medical problems that may increase the risk of micronutrient deficiencies. It is suggested that first-time mothers and, more importantly, adolescent mothers receive information related to prenatal and postnatal care from antenatal visits to healthcare providers [33]. It is important that efforts are be made to increase awareness of the unique needs and challenges faced by adolescent pregnant women, including social and economic factors that may impact their health and well-being.

### Inadequate iron folic acid supplementation (IFAS)

Folic acid is required for the rapid manufacture and growth of red blood cells which is highly needed by pregnant adolescents due to the rapid growth and expansion of blood volume and muscle mass which accompanies adolescence and pregnancy [35]. Iron-folic acid supplementation is a recommended public health strategy to pre-vent unfavourable birth outcomes and adverse hematologic complications during pregnancy [36, 37]. The WHO recommends IFAS to all pregnant women in a standard dose of 30 mg–60 mg iron and 400μg folic acid daily throughout pregnancy [38]. Bano F. [23] reported inadequate IFAS among pregnant adolescent girls. Inadequate iron-folic acid supplementation intake for pregnant adolescent women is a significant concern. These deficiencies can result in serious health problems for mothers and children. A lack of iron can lead to anaemia, which increases the risk of premature birth, low birth weight and infant mortality. Additionally, iron-folic acid deficiency in pregnancy can increase the risk of neural tube defects, endocrine disorders, and cardiac defects. Provision of IFA supplementation to adolescents from the commencement of menstruation would ensure adolescents commence pregnancy with the appropriate IFA levels. This would help prevent the reported adverse maternal and neonatal out-comes including miscarriages, stillbirths, neonatal or maternal mortality associated with inadequate intake of IFA Supplementation.

### Low maternal knowledge

Nguyen et al. [23] reported that knowledge of the benefits of consuming calcium was lower in pregnant adolescents compared to adult women. The knowledge, availability and access to essential food groups that ensure a supply of nutrients are essential in improving maternal nutrition, especially during pregnancy. Adequate maternal knowledge of the importance of micronutrients in pregnancy is crucial to the uptake of these micronutrients. Studies have shown that low maternal knowledge negatively impacts women’s health and could lead to less utilization of healthcare services and poor uptake of nutrition interventions, especially in pregnancy [39, 40]. This could also be extended to the subset of pregnant adolescent girls. A study conducted in Kenya to determine maternal iron and folic acid supplementation (IFAS) programme knowledge among pregnant women found that older women aged ≥30 years and women with higher education were more likely to have greater knowledge of the nutritional intervention programme and hence its benefits. The study also found that the level of maternal IFAS knowledge progressively increased with age and education [39]. However, there are limited studies on the impact of low maternal knowledge on micronutrient deficiency among pregnant adolescents.

### Inadequate dietary intake of micronutrients

Adequate intake of micronutrients is important for maternal health during pregnancy and optimal foetal growth and development. In the case of adolescent mothers, the long-term impacts of suboptimal nutritional status are considerable. The period of adolescence is a critical stage in human development during which lifetime behaviours are established and the dietary habits acquired during this period could enhance or undermine health throughout life [41]. Pathak et al. [25] reported a high prevalence of iron deficiency anaemia and vitamin A deficiency among pregnant adolescent mothers due to low dietary iron and vitamin A intake, respectively. Pathak et al. [25] also found that dietary nutrient intake among pregnant adolescents was significantly less compared to the recommended dietary allowance (RDA) for their respective ages and sex. With the increase in micronutrient requirements in pregnancy beyond the dietary energy requirements [42, 43], it is essential that pregnant adolescents maintain an ad-equate amount of dietary intake of micronutrients to meet up with these increased requirements. Previous studies have highlighted the need for adequate dietary intake of micronutrients by pregnant mothers to prevent adverse health outcomes in the mother and the foetus both immediately and in later life [44, 45]. This same need for adequate dietary intake could also be applicable to pregnant adolescents especially given their compromised development status.

### Strengths and Limitations

This systematic review is a comprehensive search of existing literature on micro-nutrient deficiency among pregnant adolescents in South Asia. All studies included in this review were of medium quality due to each studies’ ability to establish causal relationship between the exposure and the outcome. Despite its strengths, this study also had some limitations. First, due to the small number of studies, the generalisability, and transferability of the findings of this study to entire South Asia should be done with caution. Second, the included studies were not nation-wide studies hence caution should also be taken when generalizing individual study findings to the respective countries. Third, qualitative studies were not included in this review as the studies selected were restricted to quantitative studies. Research has shown that the inclusion of qualitative studies in systematic reviews enables triangulation of findings or offer alternative explanations [46]. Fourth, while this review summarized the underlying causes of micronutrient deficiency among pregnant adolescents, it does not address the societal causes. Finally, the discussion of the predisposing factors to micronutrient deficiency among pregnant adolescents is grossly limited due to the lack of available studies. As there is no minimum number of studies to be included in a systematic review to make it valid, the number of studies captured in this paper is largely dependent on the research topic, as well as the amount of supportive evidence available. A lack of studies reveals the field’s lack of research and knowledge gaps. However, what primarily makes a systematic review is the systematic process followed, our study adhered to PRISMA guidelines, and its protocol was duly registered with PROSPERO. It should also be noted that our study did not include grey literature. According to NIH, searching grey literature is important because sometimes only positive studies with significant findings are published in peer-reviewed literature, which can bias the results of a review. Hence, inclusion of grey literature would make for a more robust exploration of available evidence which would make our study stronger.

## Conclusion

Due to limited research, conclusions cannot be drawn about the impact of micronutrient deficiency on pregnant adolescents. The results of the present study indicate a need for more research to be undertaken in different countries of South Asia to assess the magnitude of micronutrient deficiencies amongst pregnant adolescent mothers. In this review, food insecurity and limited knowledge of adequate intake of micronutrients were key factors associated with micronutrient deficiency among pregnant adolescents. It is important to address these concerns by providing adequate health care services, education, and support to adolescent pregnant women. This can include promoting the importance of antenatal care, providing prenatal vitamins and supplements, and ensuring access to nutritious foods. Findings from this review could provide public health research with the requisite tools to establish appropriate public health interventions to minimise the burden of micronutrient deficiency among pregnant adolescents in South Asia. Integrated nutrition-sensitive interventions, including food subsidies, provision of nutrient-rich foods, and health services coupled with behaviour change, may improve the nutrition of pregnant adolescents in South Asia.

## Author Contributions

This study was designed by B.A. and K.A.; analysis was carried out by B.A., M.D. and J.B.; the manuscript was drafted by B.A. while Z.S.L., K.A and B.A. were involved in the revision and editing of the manuscript. All authors read and approved the final manuscript.

## Data Availability

All relevant data are within the manuscript and its Supporting Information files.

